# Anemia and Early-Onset Cardiovascular Risk in Premenopausal Women: Evidence from a Nationwide Cohort

**DOI:** 10.1101/2025.08.25.25334406

**Authors:** Jong-ho Park, Tae Gyun Park, Hyun Jin Choi

## Abstract

**Introduction:** Anemia, particularly iron deficiency anemia, is prevalent among reproductive-aged women and often linked to benign gynecologic disorders such as leiomyoma and adenomyosis. While anemia is a recognized contributor to hemodynamic stress, its role as an independent cardiovascular risk factor in young women remains underexplored.

**Methods:** We conducted a nationwide retrospective cohort study using Korean National Health Insurance Service data (2011–2023) including 1,706,288 premenopausal women aged 20–53 years with benign uterine disorders and no prior cardiovascular or cerebrovascular disease. Anemia was defined by ICD-10 code D50. Propensity score matching (1:1) was applied to balance cardiovascular risk factors. Outcomes included hypertension, ischemic heart disease (IHD), heart failure (HF), and cerebrovascular disease (CVD). Relative risks (RRs) were estimated overall and stratified by age group and uterine pathology.

**Results:** After matching, 627,451 women were included in each group. Anemia was associated with higher risks of hypertension (RR 1.35; 95% CI 1.34–1.37), IHD (RR 1.44; 95% CI 1.42–1.46), HF (RR 1.45; 95% CI 1.43–1.47), and CVD (RR 1.26; 95% CI 1.25–1.28). The relative risk was highest among women aged 20–29 years for hypertension (RR 1.68; 95% CI 1.41–2.02) and CVD (RR 1.45; 95% CI 1.22–1.72). Coexisting uterine leiomyoma conferred the greatest vascular risk across all outcomes.

**Conclusions:** Anemia is an independent risk factor for early-onset cardiovascular and cerebrovascular disease in women with benign uterine disorders, particularly those younger than 40 years. Incorporating gynecologic assessment into cardiovascular risk screening may enhance early prevention strategies in this population.

## Introduction

Anemia is one of the most common hematologic disorders in women of reproductive age, with iron deficiency anemia (IDA) accounting for the majority of cases worldwide(1–3). Benign uterine disorders—including leiomyoma, adenomyosis, and endometrial polyps—are frequently implicated due to their association with heavy menstrual bleeding (HMB)(4–6). Although anemia is often perceived as a gynecologic or hematologic concern, it may also be an important, yet under-recognized, cardiovascular risk factor(7–9).

From a cardiovascular standpoint, chronic anemia can lead to sustained hemodynamic stress, characterized by increased cardiac output, ventricular hypertrophy, and eventual maladaptive remodeling(10, 11). Large population-based studies have demonstrated that anemia is independently associated with increased risk of cardiovascular morbidity and mortality, even after adjustment for traditional risk factors(12–14). These findings highlight anemia as a potential early marker of vascular vulnerability, particularly in young women who may otherwise be considered at low cardiovascular risk.

Korean national epidemiologic data reveal a distinctive bimodal distribution of anemia prevalence, with peaks during adolescence and in the late reproductive years—coinciding with the highest incidence of benign uterine pathology(15, 16). Given this overlap, it is clinically relevant to investigate whether gynecologic blood loss indirectly contributes to longterm cardiovascular burden through anemia-mediated pathways.

The present study leverages the Korean National Health Insurance Service database to evaluate the association between anemia and early-onset cardiovascular disease in premenopausal women. In line with the American Heart Association’s sex-specific prevention guidelines, our findings may inform integrated strategies bridging gynecology and cardiology to improve early detection and risk reduction in women(17–20).

## Materials and Methods

A retrospective cohort study was conducted using data from the Korean National Health Insurance Service database, extracted through the National Health Information Database (NHID) for research purposes from 2011 to 2023. Initially, 14,342,579 women aged 13 to 53 years were identified. However, since the national health screening program only targets individuals aged 20 years and older, women aged 13 to 19 years were subsequently excluded from the analysis. After excluding 1,608,393 individuals aged 13-19 years and 2,905,966 with missing health screening data, a total of 9,828,220 women with at least one recorded health screening were retained. Among women with national health screening records (n = 9,828,220), we selected those with benign uterine disorders. To exclude individuals with prior cardiovascular and cerebrovascular disease, claims data from 2001 to 2010 were reviewed. Only those without a recorded diagnosis during this period were included in the final cohort, which was followed from 2011 to 2023 based on health screening records. Within this clinically defined subgroup (n = 1,706,288), we assessed the impact of anemia on vascular outcomes using propensity score matching

Of these, 399,120 were excluded due to incomplete health screening variables, and 148,381 women with a diagnosis of cardiovascular disease and cerebrovascular disease (CVD) between 2001 and 2010 were removed. The final analytic cohort included 1,706,288 women with complete data and no prior cardiovascular disease and CVD history.

Anemia was defined as a diagnosis coded as D50. Benign gynecologic diseases were defined using ICD-10 codes D25 (leiomyoma of uterus), N80 (Adenomyosis), and N84 (polyp of female genital tract). Among the final cohort, 886,323 women were classified into the anemia group and 819,965 into the non-anemia group. Propensity score matching (PSM) at a 1:1 ratio was applied based on age, body mass index (BMI), waist circumference, blood pressure, total cholesterol, triglyceride, low density lipoprotein, high density lipoprotein), aspartate aminotransferase and alanine aminotransferase , resulting in 627,451 matched pairs. Fasting blood sugar (FBS) was excluded from the matching process due to its variability. Propensity score matching was conducted using a caliper width of 0.2. Subgroup analyses were further performed by age group and by the presence of benign uterine disorders, defined by ICD-10 codes D25 (uterine leiomyoma), N80 (adenomyosis), and N84.0 (endometrial polyp).

For age-stratified analysis, participants were categorized into four groups: 20–29, 30–39, 40–49, and 50–53 years. RR values were estimated within each age group by comparing the incidence of cardiovascular outcomes between women with anemia and those with no anemia diagnosis. Additionally, to assess the impact of specific benign uterine disorders on cardiovascular risk, RR values were separately estimated within each diagnostic category using stratified models.

For each cardiovascular outcome, we calculated the risk ratio (RR) comparing disease incidence between anemic and non-anemic women across each age group. The log-transformed risk ratios (logRR) and their standard errors (SE) were computed for each age stratum.

We performed a meta-analysis using a random-effects inverse-variance model to estimate the pooled RR across all age groups for each outcome, given the observed heterogeneity. Forest plots were generated to visualize age-specific effect sizes along with their 95% confidence intervals (CI).

To assess statistical heterogeneity between age groups, we calculated the following metrics:

Tau² (between-study variance),Cochran’s Q statistic (Chi²) and corresponding p-values, and I² statistic, which quantifies the proportion of variation due to heterogeneity rather than chance.

Descriptive statistics summarized baseline characteristics, and logistic regression was employed to assess associations between anemia and vascular disease including hypertension (I10), ischemic heart disease (I20–I25), heart failure (I50), and cerebrovascular disease (I60–I69). Relative risks (RRs) were calculated by age group. Statistical significance was defined as p < 0.05. All statistical analyses were performed using SAS version 9.4 (SAS Institute Inc., Cary, NC, USA).

This study utilized publicly available and de-identified data provided by from the Korean National Health Insurance Service database, extracted through the National Health Information Database (NHID), which does not contain any personal identifying information. According to Chung-Ang University Gwangmyeong Hospital Review Board (IRB) policy and national regulations, studies using publicly available and anonymized datasets are exempt from IRB review. Therefore, IRB approval was not required for this study. The exemption status was confirmed by Chung-Ang University Gwangmyeong Hospital (IRB No. 2502-222-030).

## Results

Figure 1 presents the flowchart of study population selection. From 14,342,579 eligible women, sequential exclusions led to a final cohort of 1,706,288 women with complete variables (Figure2). Among these, 886,323 were diagnosed with anemia and 819,965 were not. After 1:1 PSM, 627,451 women remained in each group. (Figure3)

**Figure 1.**
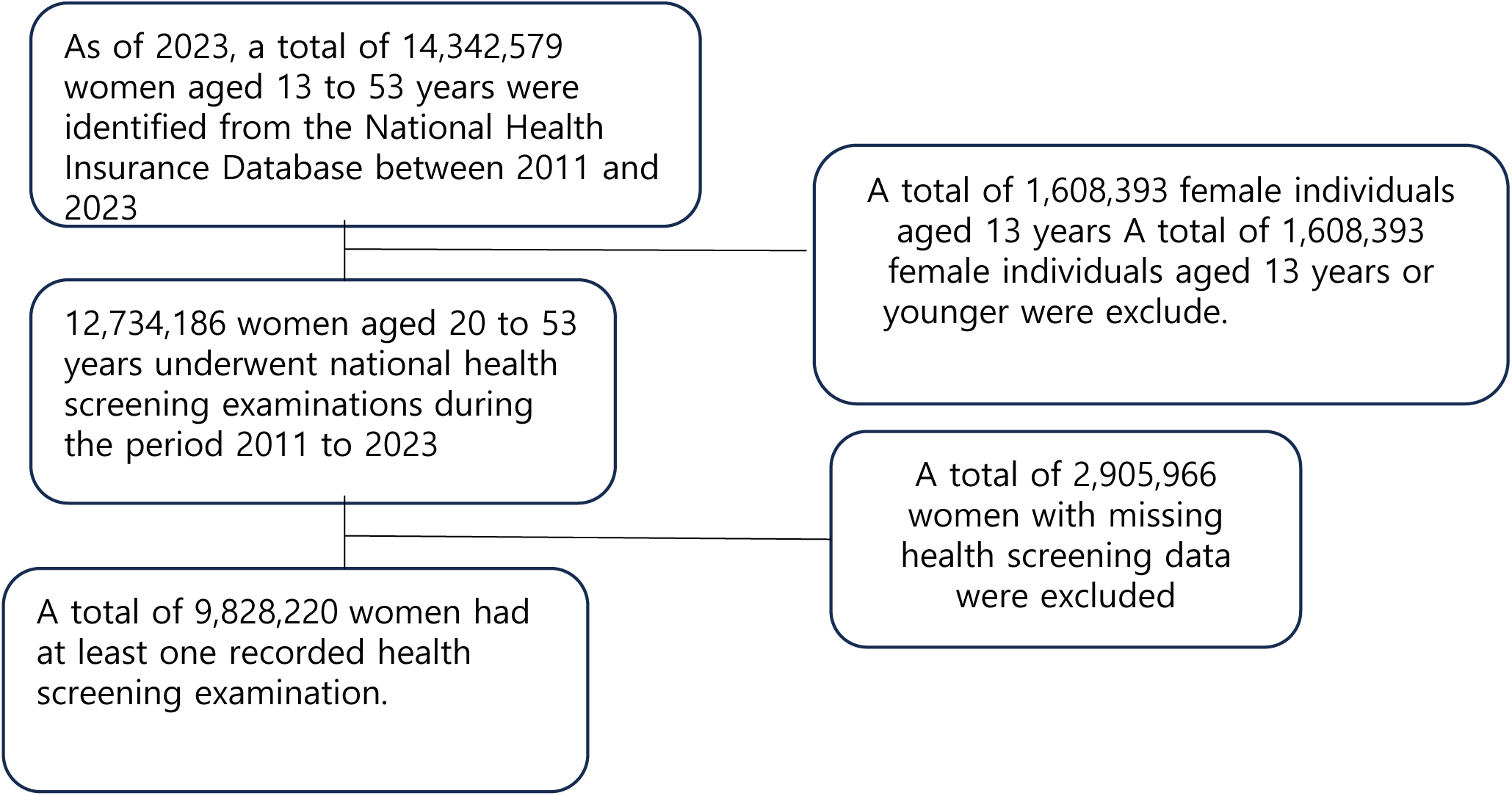
Study Population Flowchart Flow diagram depicting the inclusion and exclusion of women from the NHID cohort between 2011 and 2023. A total of 14,342,579 women aged 13–53 were initially identified. After applying exclusion criteria—such as missing health examination data or prior cardiovascular disease—a final analytic cohort of 9,828,220 women with complete health examination variables was selected.

**Figure 2.**
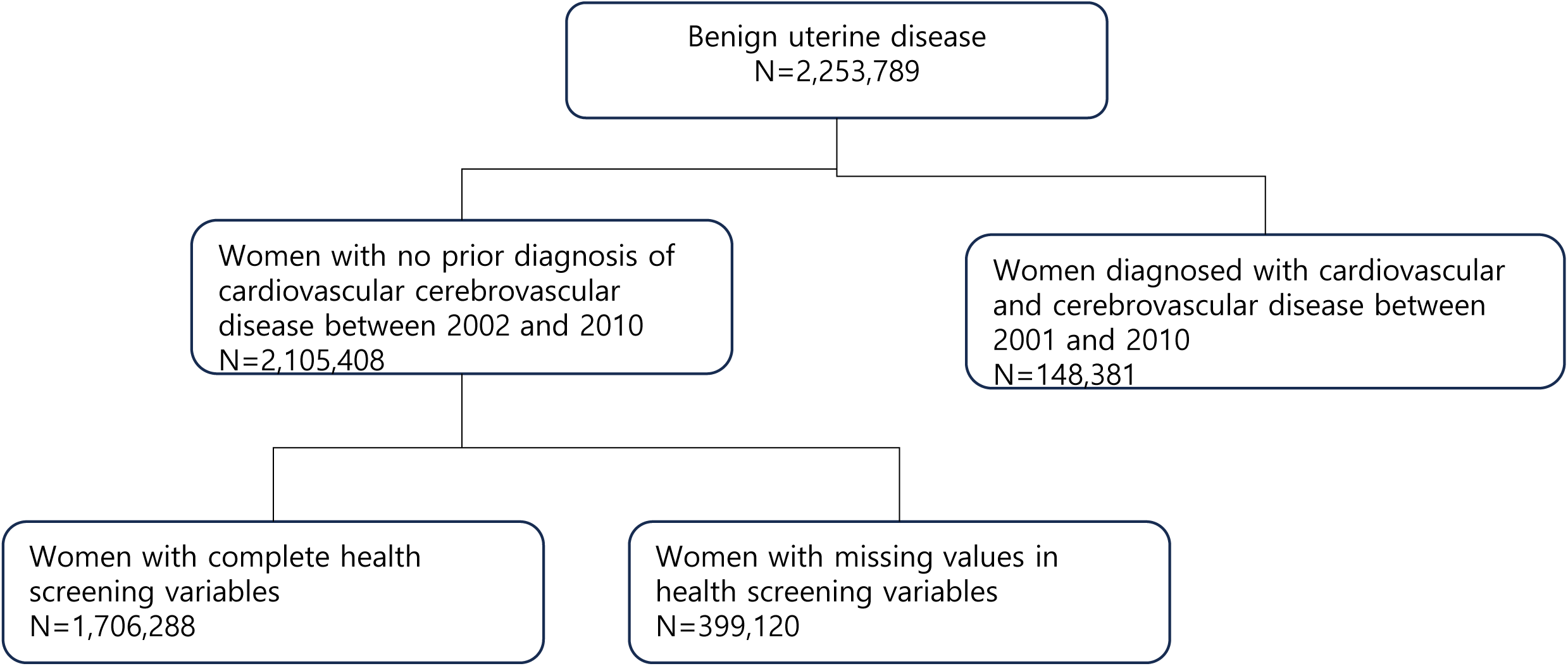
Cohort Selection Based on Cardiovascular and cerebrovascular disease History and Data Completeness History of cardiovascualr or cerebrovascular disease from 2001 to 2010 was reviewed to exclude individuals with prior disease before inclusion in the analysis cohort (2011–2023), and 148,381 were excluded. After further exclusion of women with missing health screening variables (n = 399120), a final cohort of 1,706,288 women was included in the analysis.

**Figure 3.**
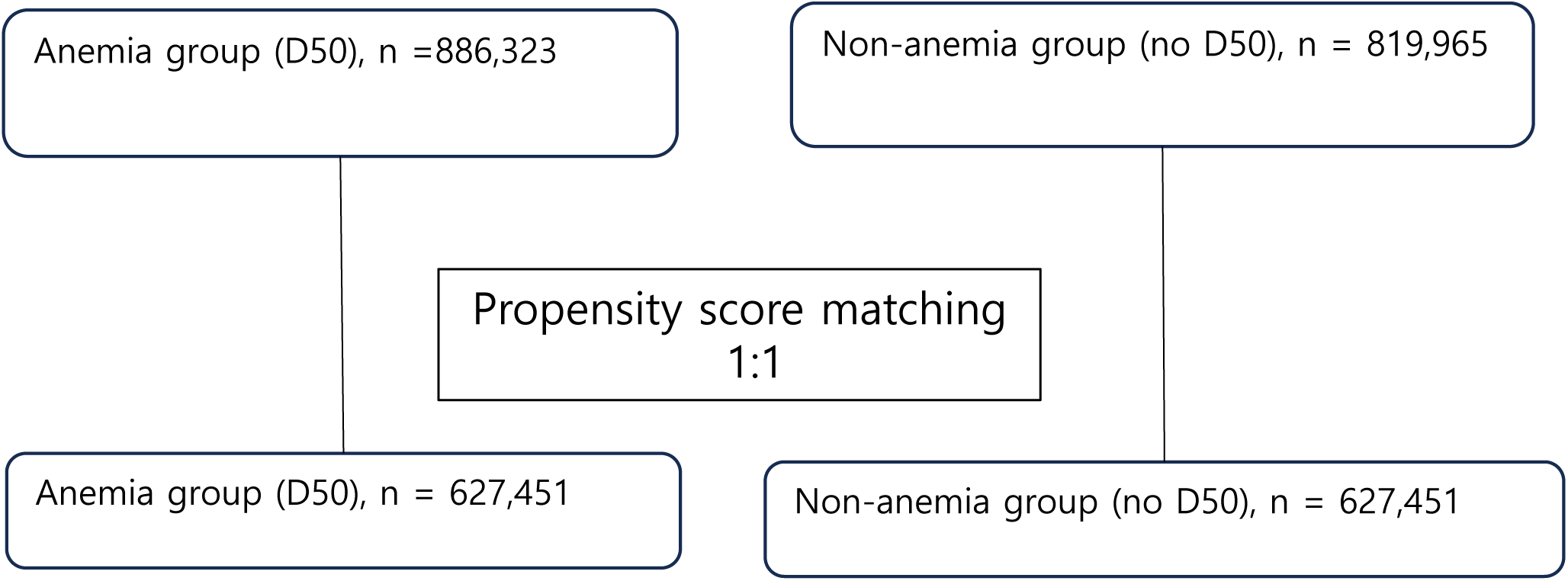
Propensity Score Matching Cohorts Distribution of study subjects before and after 1:1 propensity score matching based on age and BMI. A total of 886,323 women diagnosed with anemia (D50) and 819,965 non-anemic controls were initially identified. After matching, 627,451 women remained in each group.

Table 1 presents baseline characteristics before and after matching. Post-PSM, absolute standardized differences (ASD) for all covariates, except fasting blood sugar (FBS), were below 0.1, indicating good balance between the two groups.

**Table 1.**
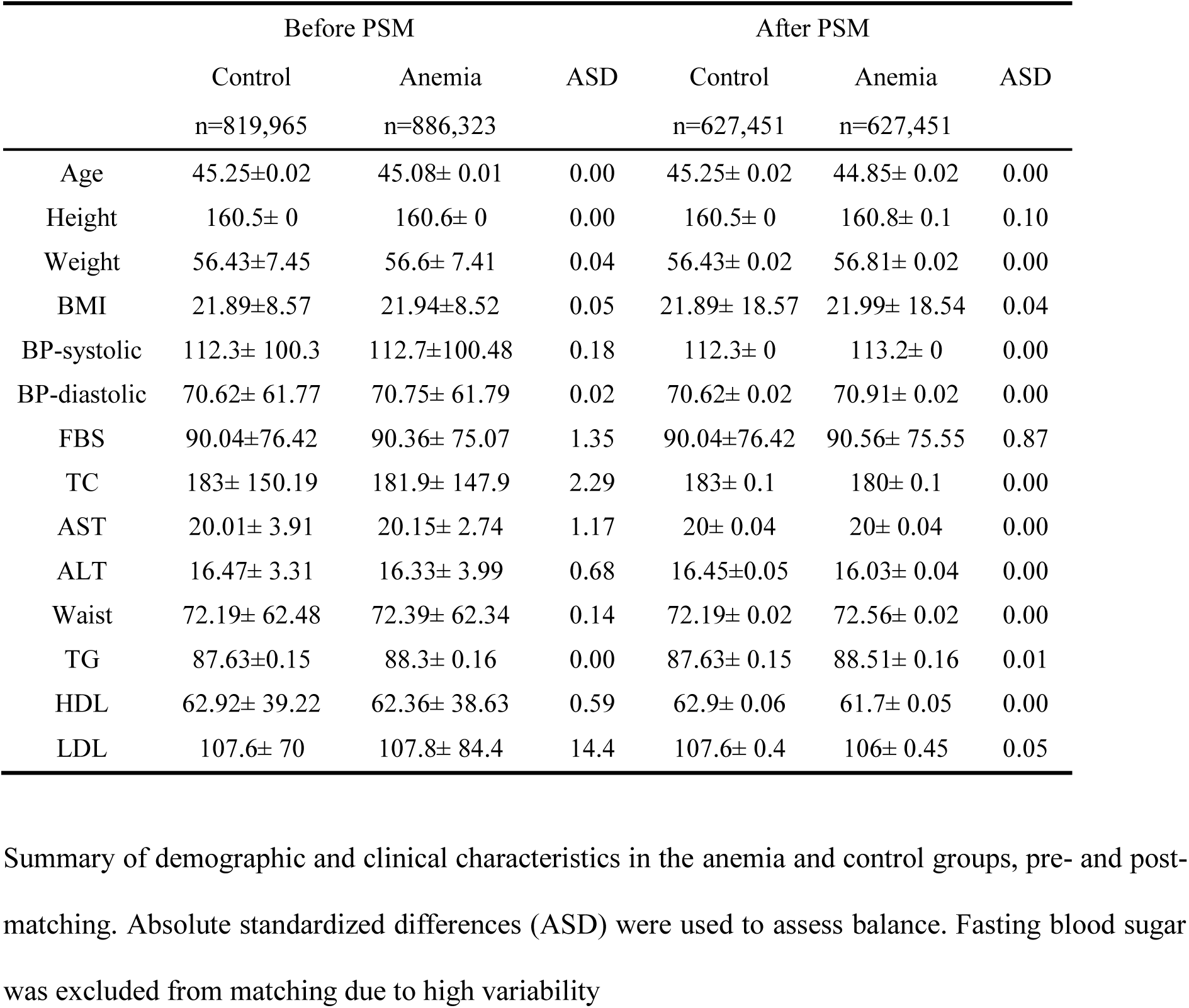
Baseline Characteristics Before and After Propensity Score Matching.

|  | Before PSM |  |  | After PSM |  |  |
| --- | --- | --- | --- | --- | --- | --- |
|  | Control | Anemia | ASD | Control | Anemia | ASD |
|  | n=819,965 | n=886,323 |  | n=627,451 | n=627,451 |  |
| Age | 45.25±0.02 | 45.08± 0.01 | 0.00 | 45.25± 0.02 | 44.85± 0.02 | 0.00 |
| Height | 160.5± 0 | 160.6± 0 | 0.00 | 160.5± 0 | 160.8± 0.1 | 0.10 |
| Weight | 56.43±7.45 | 56.6± 7.41 | 0.04 | 56.43± 0.02 | 56.81± 0.02 | 0.00 |
| BMI | 21.89±8.57 | 21.94±8.52 | 0.05 | 21.89± 18.57 | 21.99± 18.54 | 0.04 |
| BP-systolic | 112.3± 100.3 | 112.7±100.48 | 0.18 | 112.3± 0 | 113.2± 0 | 0.00 |
| BP-diastolic | 70.62± 61.77 | 70.75± 61.79 | 0.02 | 70.62± 0.02 | 70.91± 0.02 | 0.00 |
| FBS | 90.04±76.42 | 90.36± 75.07 | 1.35 | 90.04±76.42 | 90.56± 75.55 | 0.87 |
| TC | 183± 150.19 | 181.9± 147.9 | 2.29 | 183± 0.1 | 180± 0.1 | 0.00 |
| AST | 20.01± 3.91 | 20.15± 2.74 | 1.17 | 20± 0.04 | 20± 0.04 | 0.00 |
| ALT | 16.47± 3.31 | 16.33± 3.99 | 0.68 | 16.45±0.05 | 16.03± 0.04 | 0.00 |
| Waist | 72.19± 62.48 | 72.39± 62.34 | 0.14 | 72.19± 0.02 | 72.56± 0.02 | 0.00 |
| TG | 87.63±0.15 | 88.3± 0.16 | 0.00 | 87.63± 0.15 | 88.51± 0.16 | 0.01 |
| HDL | 62.92± 39.22 | 62.36± 38.63 | 0.59 | 62.9± 0.06 | 61.7± 0.05 | 0.00 |
| LDL | 107.6± 70 | 107.8± 84.4 | 14.4 | 107.6± 0.4 | 106± 0.45 | 0.05 |

Summary of demographic and clinical characteristics in the anemia and control groups, pre- and post-matching. Absolute standardized differences (ASD) were used to assess balance. Fasting blood sugar was excluded from matching due to high variability

Table 2 summarizes cardiovascular, CVD outcomes. Women with anemia had significantly elevated risks both before and after matching. Before PSM, the relative risks (RRs) were 1.35 for HTN, 1.45 for IHD, 1.45 for HF, and 1.29 for CVD. After PSM, RRs were slightly reduced but remained statistically significant: 1.35 for HTN, 1.44 for IHD, 1.45 for HF, and 1.26 for CVD. The corresponding odds ratios (ORs) before PSM were 1.43 for HTN, 1.51 for IHD, 1.51 for HF, and 1.33 for cerebral vascular disease. After PSM, the ORs were 1.44, 1.49, 1.49, and 1.29 respectively, all statistically significant (p < 0.0001).

**Table 2.**
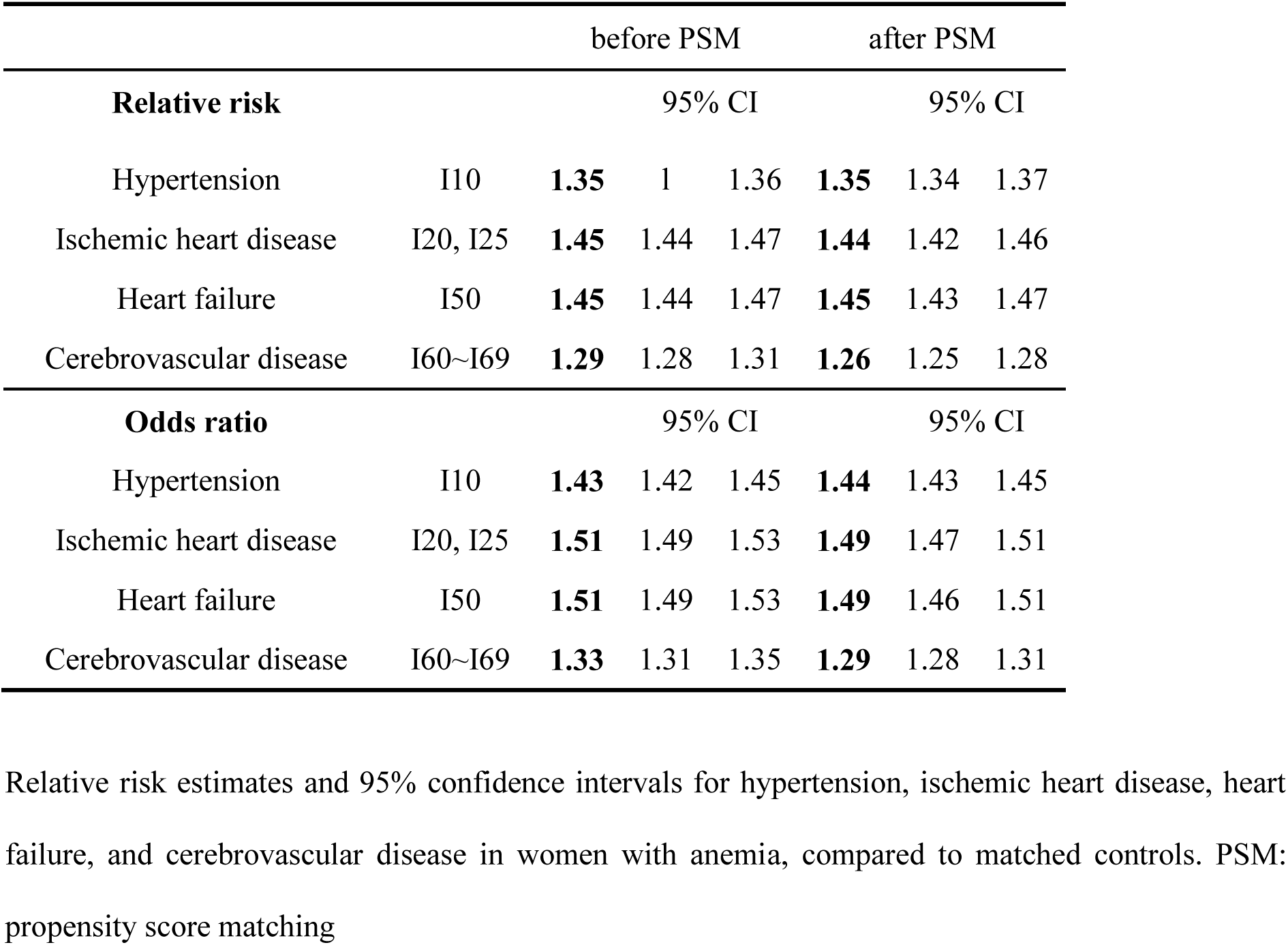
Relative Risk of Cardiovascular Outcomes in Women with Anemia.

Table 3 presents the actual number of cardiovascular disease cases stratified by age group and anemia status. The overall incidence of HTN, cardiovascular diseases and CVD increased consistently with advancing age in both the anemia and non-anemia groups. While older age groups (40–53 years) exhibited a higher absolute number of cardiovascular events, the relative proportion of disease among those with anemia was notably pronounced across all age groups, especially among younger women aged 20–39 years. For example, in women aged 30–39 years, the anemia group showed consistently higher incidence counts across all cardiovascular outcomes compared to their non-anemic cohort.

**Table 3.** Actual number of cardiovascular, cerebrovascular disease cases stratified by age and anemia status.

|  | Anemia |  |  |  |  | Non-anemia |  |  |  |  |
| --- | --- | --- | --- | --- | --- | --- | --- | --- | --- | --- |
|  | n=627,451 |  |  |  |  | n=627,451 |  |  |  |  |
|  | Age | 20~29 | 30~39 | 40~49 | 50~53 | Age | 20~29 | 30~39 | 40~49 | 50~53 |
|  | Age count | 5,502 | 132,179 | 303,754 | 186,016 | Age count | 4,384 | 119,052 | 305,290 | 198,725 |

|  |  | Rate | 0.88% | 21.07% | 48.41% | 29.65% | Rate | 0.70% | 18.97% | 48.66% | 31.67% |
| --- | --- | --- | --- | --- | --- | --- | --- | --- | --- | --- | --- |
| Number of patients | **HTN** | 123,350 | 390 | 13,301 | 56,671 | 52,988 | 91,070 | 184 | 8,013 | 40,623 | 42,250 |
|  | **IHD** | 63,115 | 419 | 9,979 | 29,872 | 22,845 | 43,779 | 218 | 5,762 | 20,077 | 17,722 |
|  | **HF** | 46,851 | 345 | 7,886 | 22,230 | 16,390 | 32,330 | 177 | 4,708 | 15,025 | 12,420 |
|  | **CVD** | 65,995 | 381 | 8,866 | 30,744 | 26,004 | 52,295 | 210 | 5,744 | 23,694 | 22,647 |

HTN: Hypertension, IHD: Ischemic heart disease, HF: Heart failure, CVD: Cerebrovascular disease

Table 4 demonstrates the relative risks (RRs) of cardiovascular diseases associated with anemia across different age groups. Although absolute disease incidence increased with age, the RR as-sociated with anemia was inversely related to age, being highest in the youngest groups. Specifically, women in their 20s with anemia had markedly increased relative risks for HTN, IHD, HF, and CVD compared to older age groups. The highest RR was observed for HTN among women aged 20–29 years (RR 1.68, 95% CI 1.41–2.02), and this RR progressively decreased with advancing age, indicating that the impact of anemia as an independent cardiovascular risk factor is particularly significant among younger women.

**Table 4.** Age-Stratified Relative Risk of Cardiovascular Disease.

| Age_group(yrs) | Disease | Relative risk | 95% CI |  |
| --- | --- | --- | --- | --- |
| 20~29 | Hypertension | 1.6845 | 1.4073 | 2.0164 |
|  | Ischemic heart disease | 1.5315 | 1.2941 | 1.8124 |
|  | Heart failure | 1.5531 | 1.2901 | 1.8696 |
|  | Cerebrovascular disease | 1.4456 | 1.2159 | 1.7188 |
| 30~39 | Hypertension | 1.4951 | 1.4526 | 1.5388 |
|  | Ischemic heart disease | 1.5599 | 1.5087 | 1.6128 |
|  | Heart failure | 1.5087 | 1.454 | 1.5654 |
|  | Cerebrovascular disease | 1.3902 | 1.3436 | 1.4385 |
| 40~49 | Hypertension | 1.4022 | 1.3831 | 1.4215 |
|  | Ischemic heart disease | 1.4954 | 1.4679 | 1.5234 |
|  | Heart failure | 1.487 | 1.4557 | 1.519 |
|  | Cerebrovascular disease | 1.3041 | 1.2813 | 1.3274 |
| 50~53 | Hypertension | 1.3398 | 1.3209 | 1.3591 |
|  | Ischemic heart disease | 1.5858 | 1.5523 | 1.6201 |
|  | Heart failure | 1.4098 | 1.3762 | 1.4443 |
|  | Cerebrovascular disease | 1.2267 | 1.2037 | 1.2501 |

Relative risk of cardiovascular diseases according to age group. Risk of hypertension was highest in women in their 20s with anemia, and progressively decreased with age

Figure 4 visualizes the trends of relative risk (RR) across different disease categories by age group. For most conditions examined—Ischemic heart disease(IHD), CVD, HF, and HTN women in their 30s exhibited higher risk estimates (CVD: 1.39, HF: 1.51, HTN: 1.50) compared to women in their 40s (CVD: 1.30, HF: 1.49, HTN: 1.40) and 50s (CVD: 1.23, HF: 1.41, HTN: 1.34). However, for IHD, the highest risk was observed in women in their 50s (RR 1.59 [1.55; 1.62]).

**Figure 4.**
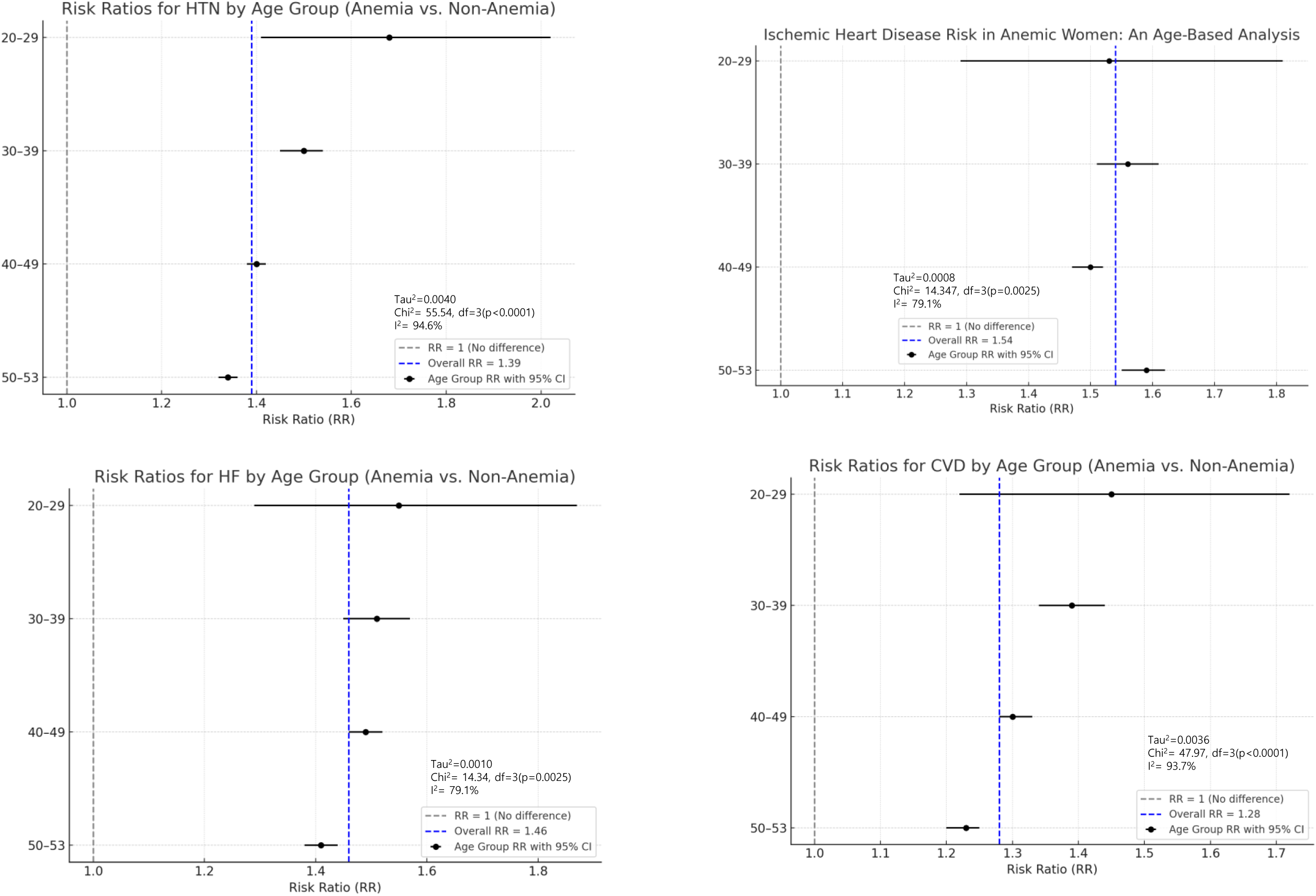
Age-Stratified Relative Risk of Vascular Diseases Associated with Anemia in Premenopausal Women. Forest plots show relative risks (RRs) and 95% confidence intervals for cardiovascular outcomes across age groups. Anemia was associated with higher RRs in younger women for CVD, HF, and HTN, while the highest RR for IHD was observed in women in their 50s. Heterogeneity metrics (Tau², Chi², I²) indicate significant between-group variation.

Although point estimates were highest among women in their 20s for CVD (1.45), HF (1.55), and HTN (1.68), their wider confidence intervals (CVD: [1.22; 1.72], HF: [1.29; 1.87], HTN: [1.41; 2.02]) suggest lower statistical precision, likely attributable to smaller case numbers. Based on these point estimates, the anemia-associated risks for HTN and CVD were highest in women in their 30s, whereas the risk for IHD peaked among women in their 50s.

Subgroup analysis by type of benign uterine disease (Table 5) showed that women with uterine leiomyoma (D25) had the highest relative risks for cardiovascular outcomes, including HTN (RR 1.84; 95% CI 1.83–1.85), IHD (RR 1.75; 95% CI 1.74–1.77), HF (RR 1.75; 95% CI 1.72–1.77), and CVD (RR 1.83; 95% CI 1.82–1.85). Although cardiovascular risks were also significantly elevated in women with adenomyosis (N80) and endometrial polyps (N84), the magnitude of risk was relatively lower compared to those with uterine leiomyoma.

**Table 5.** Relative Risk (RR) of Vascular Disease by Type of Benign Uterine Disease.

| ICD-10 code | Diagnosis | RR | 95% CI |  |
| --- | --- | --- | --- | --- |
| D25<br>(Uterine myoma) | Hypertension | 1.8411 | 1.831 | 1.8512 |
|  | Ischemic heart disease | 1.7501 | 1.7351 | 1.7681 |
|  | Heart failure | 1.7461 | 1.7244 | 1.7681 |
|  | Cerebrovascular disease | 1.8326 | 1.8158 | 1.8495 |
| N840<br>(Uterine polyp) | Hypertension | 1.4011 | 1.3917 | 1.4106 |
|  | Ischemic heart disease | 1.4714 | 1.4564 | 1.4866 |
|  | Heart failure | 1.465 | 1.4433 | 1.487 |
|  | Cerebrovascular disease | 1.4352 | 1.4191 | 1.4514 |
| N80<br>(Adenomyosis) | Hypertension | 1.7539 | 1.7442 | 1.7637 |
|  | Ischemic heart disease | 1.7761 | 1.761 | 1.7914 |
|  | Heart failure | 1.8276 | 1.8054 | 1.8502 |
|  | Cerebrovascular disease | 1.8071 | 1.7905 | 1.8239 |

## Discussion

### Principal Findings

In this nationwide cohort study including more than 1.7 million premenopausal women with benign uterine disorders, we found that anemia was independently associated with a significantly higher risk of cardiovascular and cerebrovascular disease. After rigorous propensity score matching, women with anemia showed increased risks of hypertension, ischemic heart disease, heart failure, and cerebrovascular disease compared with women without anemia. The excess risk was most pronounced in women in their 20s and 30s, suggesting that anemia may represent an early marker of vascular vulnerability in reproductive-aged women. Although the absolute number of cardiovascular events was greater among older women, the relative contribution of anemia to vascular risk was consistently elevated in younger age groups.

### Comparison with Existing Literature

Our findings expand upon prior studies linking anemia to adverse cardiovascular outcomes. Earlier investigations in patients with chronic kidney disease and heart failure have consistently demonstrated that anemia worsens prognosis and accelerates disease progression(21, 22). At the population level, observational studies such as the ARIC cohort reported that anemia was associated with a higher incidence of cardiovascular disease, but these studies rarely focused on younger women(23). The present study addresses this gap by demonstrating that anemia, particularly in the context of benign gynecologic disorders, independently elevates vascular risk at a relatively early stage of life.

Benign uterine disorders, including leiomyoma, adenomyosis, and endometrial polyps, are common causes of heavy menstrual bleeding and the leading contributors to iron deficiency anemia in premenopausal women(4, 6). Previous cohort studies have shown that women with leiomyomas have increased hospitalization rates for cardiovascular conditions and related comorbidities(24). Our results build on these observations by explicitly linking gynecologic disease, anemia, and subsequent cardiovascular outcomes within a unified analytic framework.

### Mechanistic Considerations

The biological plausibility of this association is supported by several mechanisms. Chronic anemia reduces oxygen-carrying capacity, leading to compensatory increases in cardiac output and left ventricular hypertrophy. Persistent hemodynamic stress may predispose to myocardial dysfunction and endothelial injury(8). Experimental studies also suggest that iron deficiency impairs mitochondrial function and increases oxidative stress, further promoting vascular dysfunction. In the cerebral circulation, chronic anemia alters blood viscosity and cerebral blood flow, potentially increasing susceptibility to ischemic injury(10). Taken together, these mechanisms provide a coherent explanation for the increased cardiovascular and cerebrovascular risks observed in our study.

### Hypertension and Cerebrovascular Disease as Early Manifestations

Among the cardiovascular outcomes examined, the relative risks for hypertension and cerebrovascular disease were particularly elevated in younger women. Hypertension is a well-recognized precursor of later cardiovascular morbidity, and early-onset hypertension carries a disproportionately high long-term burden. The finding that women in their 20s with anemia had nearly 70% higher risk of developing hypertension emphasizes the potential role of anemia as an early-life determinant of vascular health. Similarly, the excess risk of stroke observed in young women with anemia suggests that cerebrovascular vulnerability may manifest earlier than expected, highlighting the need for timely recognition and intervention.

### Interpretation of Hysterectomy-Related Risk

A recent nationwide Korean study reported that hysterectomy performed in women aged 40–49 years was associated with increased cardiovascular risk, particularly stroke(20). Our findings provide context for interpreting these results. It is likely that many of the women who underwent hysterectomy had already experienced prolonged symptomatic gynecologic disease and anemia before surgery. Thus, the elevated risk observed after hysterectomy may reflect both the surgical event and cumulative vascular damage from longstanding anemia. This distinction underscores the importance of evaluating cardiovascular outcomes in relation not only to surgical interventions but also to the underlying trajectory of gynecologic disease and hematologic burden. Future prospective studies should investigate whether early intervention for heavy menstrual bleeding— before hysterectomy becomes necessary—can mitigate long-term cardiovascular risk.

### Clinical Implications

The clinical implications of our findings are substantial. Routine screening for anemia in reproductive-aged women, particularly those with benign gynecologic disorders, may provide an opportunity for early cardiovascular prevention. Interventions such as iron supplementation, hormonal therapy, and minimally invasive gynecologic procedures not only improve quality of life but may also reduce downstream vascular complications. This approach is consistent with the American Heart Association’s sex-specific prevention guidelines, which emphasize the importance of incorporating reproductive and hematologic factors into cardiovascular risk assessment in women(25).

Moreover, the results highlight the need for closer collaboration between gynecology and cardiology. Women presenting with abnormal uterine bleeding should undergo not only gynecologic evaluation but also cardiovascular risk screening. Conversely, young women presenting with hypertension or cerebrovascular events of unclear etiology should be evaluated for possible anemia or gynecologic conditions. Such an integrated approach has the potential to improve early detection and prevention of cardiovascular disease in women.

### Public Health and Economic Implications

From a public health perspective, the implications of untreated anemia extend beyond individual morbidity. Cardiovascular disease is the leading cause of mortality in women worldwide, and its prevalence is increasing in many regions(26). The Global Burden of Disease study identified iron deficiency anemia as the most common nutritional disorder globally, disproportionately affecting women of reproductive age(27). Addressing anemia in this population may therefore reduce both gynecologic morbidity and long-term cardiovascular disease burden.

Economic analyses have demonstrated that definitive treatments for refractory uterine bleeding, such as hysterectomy, are cost-effective when long-term healthcare utilization is considered(28). However, our findings suggest that earlier intervention may yield even greater benefits. By preventing progression to severe anemia and reducing future cardiovascular events, proactive management of benign uterine disorders may improve both patient-centered outcomes and overall healthcare efficiency.

### Strengths and Limitations

The major strengths of this study include the use of a large, nationally representative cohort, robust propensity score matching, and age-stratified analyses that provide clinically relevant insights into the timing of risk. The size and scope of the database allowed for detailed evaluation of associations across a wide age spectrum and multiple cardiovascular outcomes.

Nevertheless, several limitations must be acknowledged. Anemia was defined by ICD-10 codes rather than laboratory values, limiting our ability to evaluate severity or treatment response. Residual confounding from unmeasured factors such as smoking, body mass index, and physical activity cannot be excluded. Additionally, our focus on women with benign uterine disorders means that we did not examine the independent effect of gynecologic disease without anemia.

Finally, as with any retrospective study using administrative data, the potential for misclassification bias exists.

### Future Directions

Future research should determine whether correction of anemia in women with benign gynecologic disorders can alter cardiovascular outcomes. Randomized trials evaluating the impact of iron therapy, hormonal interventions, or surgical management on long-term vascular events would provide critical causal evidence. Integration of laboratory data, imaging studies, and biomarkers of vascular health may clarify mechanistic pathways. Moreover, international studies across diverse populations are needed to confirm the generalizability of these findings beyond the Korean context.

## Conclusion

In conclusion, anemia in women with benign uterine disorders is an independent risk factor for early-onset cardiovascular and cerebrovascular disease. The excess risk is particularly pronounced in younger women, suggesting that anemia may serve as an early marker of vascular vulnerability. Early recognition and integrated gynecologic–cardiovascular management may reduce both gynecologic symptom burden and long-term vascular morbidity, offering significant clinical and public health benefits.

## Data Availability

The National Health Insurance Service's dataset is not permitted to be taken outside. (If it is taken outside, you can be criminally punished under the Personal Information Protection Act.) Therefore, you must select NON APPLICABLE

## Declaration of generative AI and AI-assisted technologies in the writing process

During the preparation of this work the author(s) used [ChatGPT-4o,5 (Scholar GPT)] in order to for purposes including formatting, English editing. After using this tool/service, the author(s) reviewed and edited the content as needed and take(s) full responsibility for the content of the publication

